# Lack of antibodies against seasonal coronavirus OC43 nucleocapsid protein identifies patients at risk of critical COVID-19

**DOI:** 10.1101/2020.12.07.20245241

**Authors:** Martin Dugas, Tanja Grote-Westrick, Uta Merle, Michaela Fontenay, Andreas E. Kremer, Frank Hanses, Richard Vollenberg, Eva Lorentzen, Shilpa Tiwari-Heckler, Jérôme Duchemin, Syrine Ellouze, Marcel Vetter, Julia Fürst, Philipp Schuster, Tobias Brix, Claudia M. Denkinger, Carsten Müller-Tidow, Hartmut Schmidt, Phil-Robin Tepasse, Joachim Kühn

## Abstract

Most COVID-19 patients experience a mild disease; a minority suffers from critical disease.

We report about a biomarker validation study regarding 296 patients with confirmed SARS-CoV-2 infections from four tertiary care referral centers in Germany and France.

Patients with critical disease had significantly less anti-HCoV OC43 nucleocapsid protein antibodies compared to other COVID-19 patients (p=0.007). In multivariate analysis, OC43 negative inpatients had an increased risk of critical disease, higher than the risk by increased age or BMI, and lower than the risk by male sex. A risk stratification based on sex and OC43 serostatus was derived from this analysis.

Our results indicate that prior infections with seasonal human coronaviruses can protect against a severe course of COVID-19. Anti-OC43 antibodies should be measured for COVID-19 inpatients and considered as part of the risk assessment. We expect individuals tested negative for anti-OC43 antibodies to particularly benefit from vaccination, especially with other risk factors prevailing.

## Introduction

Approximately 10 to 20 percent of COVID-19 patients require hospital treatment and about a quarter of those need to be treated in intensive care units (ICU) (1). In contrast, the majority of COVID-19 patients experience mild symptoms only. Known important risk factors are age, male sex, high body mass index and pre-existing comorbidities (2). However, severe or fatal COVID-19 occurs also in seemingly healthy individuals without obvious risk factors. COVID-19 disease heterogeneity is still not well understood and complicates patient care.

There are several reports that cross-protection against SARS-CoV-2 might contribute to this phenomenon; for example, Grifoni (3), Le Bert (4) and Mateus (5) have described T cell responses to SARS-CoV-2 in unexposed human individuals. In a recent manuscript, Henss (6) stated that disease severity seemed to correlate with low NL63-neutralizing activities, suggesting the possibility of cross-reactive protection. Shrock (7) found that that patients who had required hospitalization exhibited stronger and broader antibody responses to SARS-CoV-2 but weaker overall responses to past infections compared with those who did not need hospitalization. In a survey of 1,186 convalescent patients with mild COVID-19 (8), contact to small children has been reported frequently (30.1% of participants). Potentially, childhood-related infections might modify disease severity of COVID-19 and thereby contribute to the low incidence of severe infections in small children (9). Infections with human coronaviruses (HCoV) NL63, 229E, HKU1 and OC43 frequently occur in children and are a typical cause of acute infections of the upper respiratory tract during the cold season.

A pilot study with 60 patients at University Hospital Münster reported less severe course of COVID-19 in patients with elevated levels of antibodies against seasonal human coronaviruses (sHCoV) OC43 and HKU1 (10; preprint under review), which like SARS-CoV-2 belong to the genus *betacoronavirus* of the subfamily *orthocoronaviridae*. To validate the findings from the pilot study, a study with an independent patient cohort was conducted according to the same protocol.

## Results

### Presence of NP-specific IgG antibodies against sHCoVs

Seroreactivity against sHCoVs was tested in a total of 296 human sera. Details of patient cohort are given in materials and methods. No such antibodies (-) were measured for OC43 in 22%, for HKU1 in 18%, for NL63 in 14% and for NP229E in 17% of cases. Antibody level below (+/-) or with cutoff (+) was found for OC43 in 64%, for HKU1 in 50%, for NL63 in 44% and for NP229E in 48% of patients. NP-specific IgG antibodies (++/+++) were detected for OC43 in 14%, for HKU1 in 32%, for NL63 in 43% and for NP229E in 35% of patients, respectively.

### sHCoV-specific IgG antibody levels in relation to symptom onset

We assessed sHCoV-specific IgG antibody levels in relation to symptom onset (days after start of COVID-19 symptoms) for the validation cohort. Figure 1 presents results from this analysis: We observed no systematic effect of the time point of sample collection on relative IgG antibody levels against HCoV OC43 and HKU1 NP. However, regarding HCoV NL63 and 229E, antibody levels were significantly correlated with time since symptom onset, i.e., increasing HCoV IgG antibody levels over time were determined for NL63 (p=0.0017) and 229E (p=0.00018). Median time point of sample collection was 8 days after symptom onset for inpatients and 5 days for outpatients (p<0.0001).

**Figure 1:**
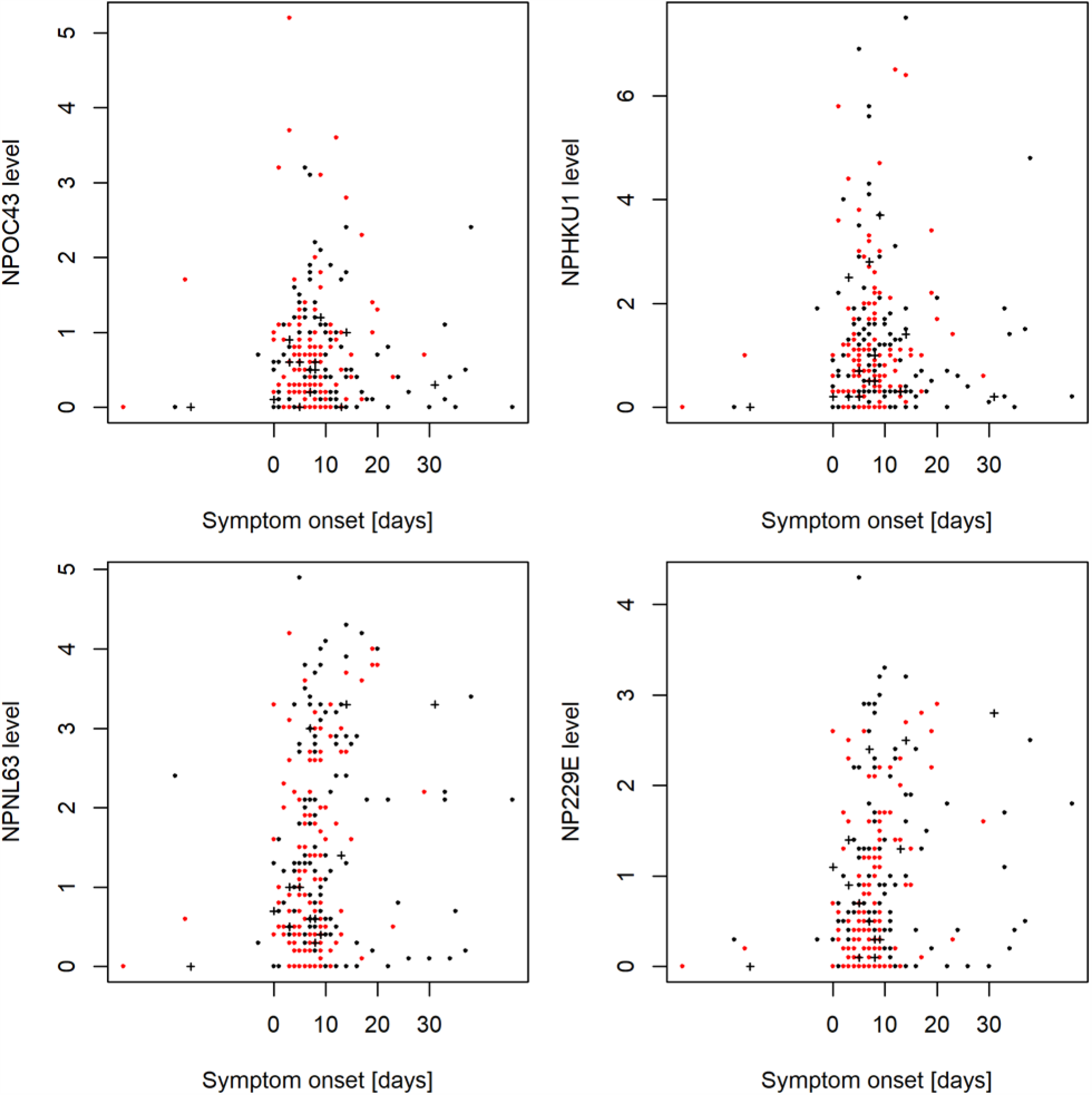
sHCoV IgG antibody levels for the validation cohort at different time points during SARS-CoV-2 infection. Black dots indicate male patients, red dots female patients. Crosses denote fatal cases. Relative IgG antibody levels against NP of sHCoVs were determined by immunostrip assay as given in Materials and Methods. No association was identified for OC43 (p=0.73) and HKU1 (p=0.89). Increasing HCoV IgG antibody levels over time were determined for NL63 (p=0.0017) and 229E (p=0.00018). In 5 patients serum collected before the SARS-CoV-2 infection was available.

### Risk of critical COVID-19 for inpatients in relation to HCoV OC43 serostatus

Table 1 presents detailed patient counts for the validation cohort regarding HCoV OC43 serostatus and type of treatment. Frequency of critical disease in COVID-19 inpatients was significantly different according to OC43 status (p=0.014). OC43 negative inpatients required ICU therapy more frequently; thus the key result from the pilot study was validated.

**Table 1:** Disease severity of COVID-19 inpatients by HCoV OC43 serostatus in the validation cohort. HCoV OC43-specific IgG antibody levels were rated on an ordinal scale as given in Materials and Methods: non-detectable (-), below cutoff (+/-), with cutoff intensity (+), above cutoff (++), and very strong intensity (+++). 70% of OC43 negative inpatients developed critical disease (treatment on ICU). In contrast, 35% of inpatients with high levels of anti-OC43 antibodies required ICU therapy (p=0.014).

|  |  |  |
| --- | --- | --- |
| 296 COVID patients |  |  |
| 106 outpatients |  |  |
| 190 inpatients | 44 OC43 negative (-) | 70% ICU<br>31 ICU<br>13 non-ICU |
|  | 120 OC43 below (+/-) or with cutoff intensity (+) | 55% ICU<br>66 ICU<br>54 non-ICU |
|  | 26 OC43 above cutoff intensity (++) or very strong intensity (+++) | 35% ICU<br>9 ICU<br>17 non-ICU |

Figure 2 presents levels of sHCoV-specific IgG antibody levels on an ordinal scale for inpatients (ICU group compared to non-ICU group; without outpatients). Patients with critical disease had significantly less anti-HCoV OC43 nucleocapsid protein antibodies compared to other COVID-19 inpatients (p=0.007). In more detail, the proportion of patients without sHCoV antibodies (“-”) was increased for critical disease. This pattern was most pronounced for HCoV OC43: odds ratio (OR) 2.26 [95% CI 1.09 - 4.66], followed by HCoV NP229E (OR 1.57 [95% CI 0.73 - 3.40]), HCoV HKU1 (OR 1.14 [95% CI 0.55 - 2.37]) and HcoV NL63 (OR 1.45 [95% CI 0.58 - 3.63]). Vice versa, patients without critical disease presented antibody levels above cutoff (++) or very strong antibody reactivities (+++) against sHCoV more frequently.

**Figure 2:**
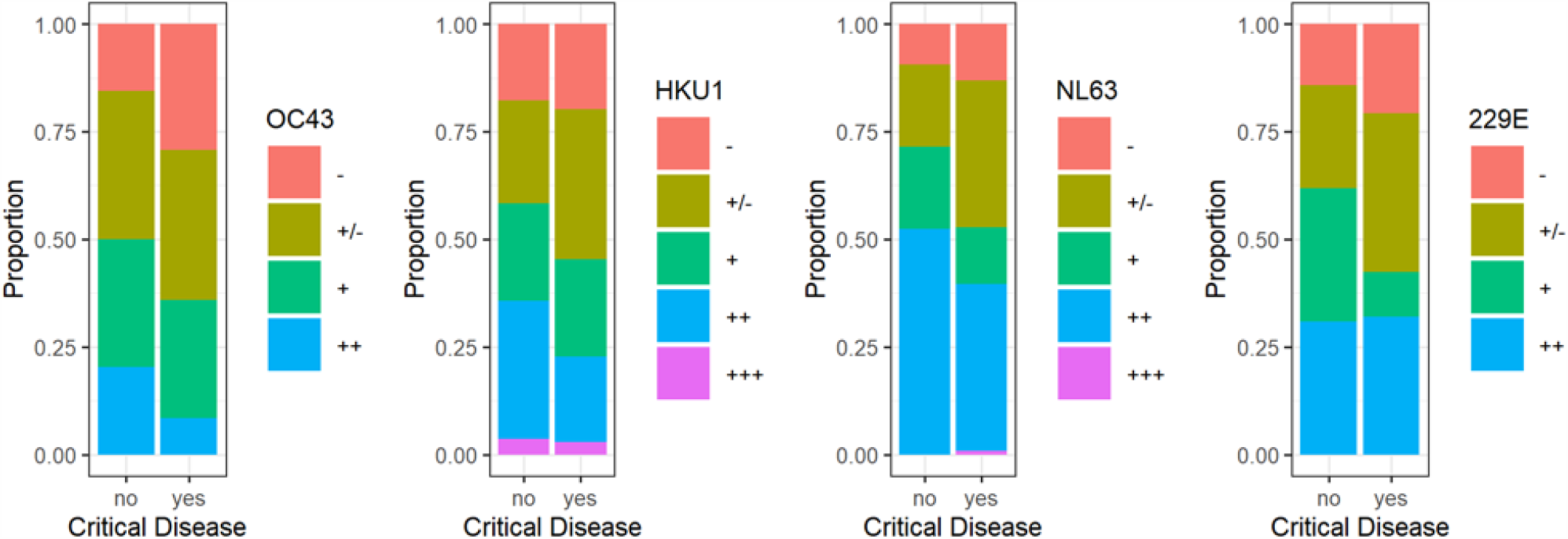
Proportion of ordinal sHCoV antibody levels from COVID-19 inpatients with and without critical disease. Patients with critical disease presented non-detectable antibody levels more frequently than inpatients without critical disease. This difference was most pronounced for HCoV OC43 (odds ratio 2.26 [95% CI 1.09 - 4.66]). Patients without critical disease presented antibody reactivities above cutoff (++) or very strong reactivities (+++) against sHCoV more frequently. Again, this difference was most pronounced for HCoV OC43 (odds ratio 2.73 [95% CI 1.15 - 6.50]).

### OC43 serostatus as independent risk factor

Sex, age and BMI are known risk factors for COVID-19 severity. Thus, the association between HCoV OC43 antibody levels and these risk factors was analyzed (supplemental figure 3). There was no significant difference in HCoV OC43 antibody levels between males and females in the validation cohort (p=0.56). There was no significant association between HCoV OC43 antibody levels and age (p=0.07) or BMI (p=0.88). Therefore, HCoV OC43 antibody levels may provide independent information in addition to known risk factors. Figure 3 presents rate of ICU therapy by sex, OC43 antibody level, age and BMI.

**Figure 3:**
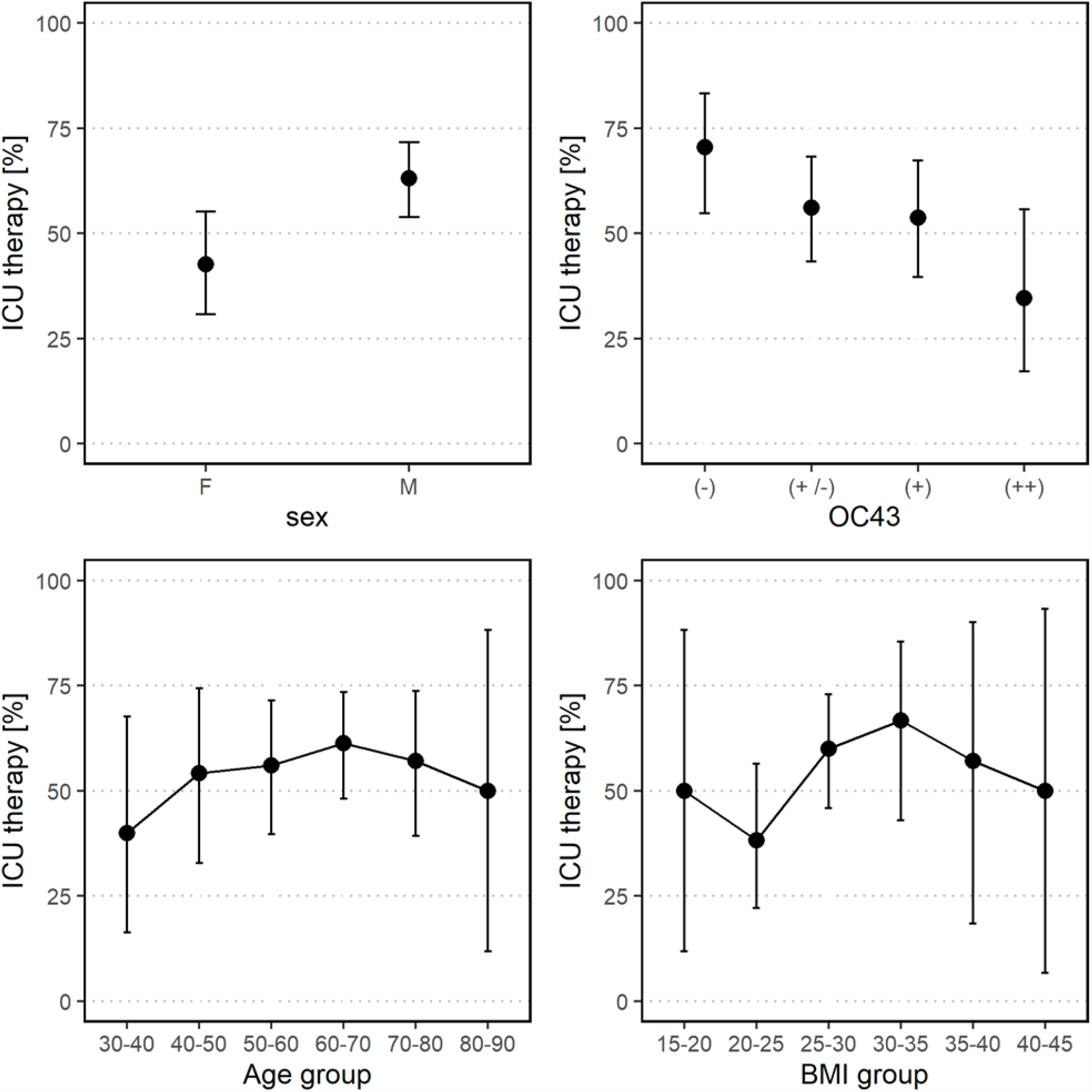
Rate of ICU therapy by sex, OC43 serostatus, age and BMI group regarding COVID-19 inpatients. Rates are provided with 95% confidence intervals. Males and OC43 negative patients require ICU therapy most frequently.

We performed multivariate analysis of inpatients to compare the effect size of OC43 with established risk factors age, sex and BMI. In binary logistic regression, absence of HCoV OC43-specific antibodies (adjusted odds ratio (AOR) 2.68 [95% CI 1.09 - 7.05]) and male sex (AOR 2.98 [95% CI 1.38 - 6.63]) were the strongest predictors for critical disease, while age (AOR 1.02 [95% CI 0.99 - 1.04]) and BMI (AOR 1.07 [95% CI 1.00 - 1.16]) did not reach statistical significance in this cohort. It has to be considered that median age of inpatients in our validation cohort was 61 years. Table 2 presents a risk stratification of COVID-19 inpatients by sex and OC43 serostatus derived from this multivariate analysis.

**Table 2:**
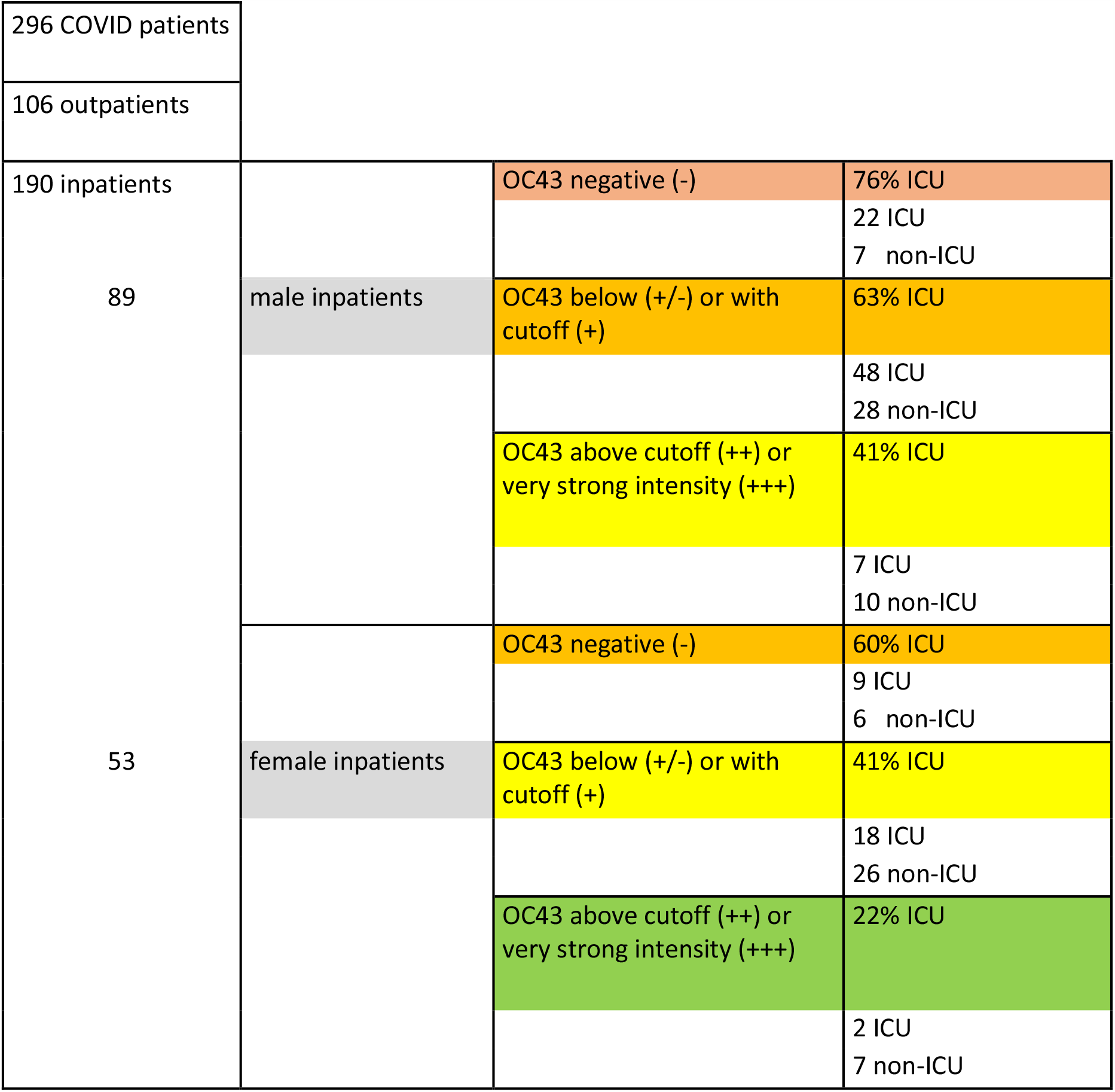
COVID-19 inpatient risk stratification by sex and OC43 serostatus. Multivariate analysis identified sex and OC43 serostatus as strongest predictors for critical disease. Age and BMI did not reach statistical significance in this cohort. 90% of males in this cohort were 42 years or older; 90% of females in this cohort were 38 years or older. For this reason, this table can be applied for inpatients aged 40 years or older to estimate risk of critical disease. For example, an OC43 negative female inpatient would have an estimated risk of 60%.

### Length of stay and sHCoV-specific IgG antibody levels

Figure 4 presents hospital length of stay (LoS) in relation to IgG antibody levels against sHCoV. Of note, a combination of high sHCoV antibody levels and long LoS occurred infrequently.

**Figure 4:**
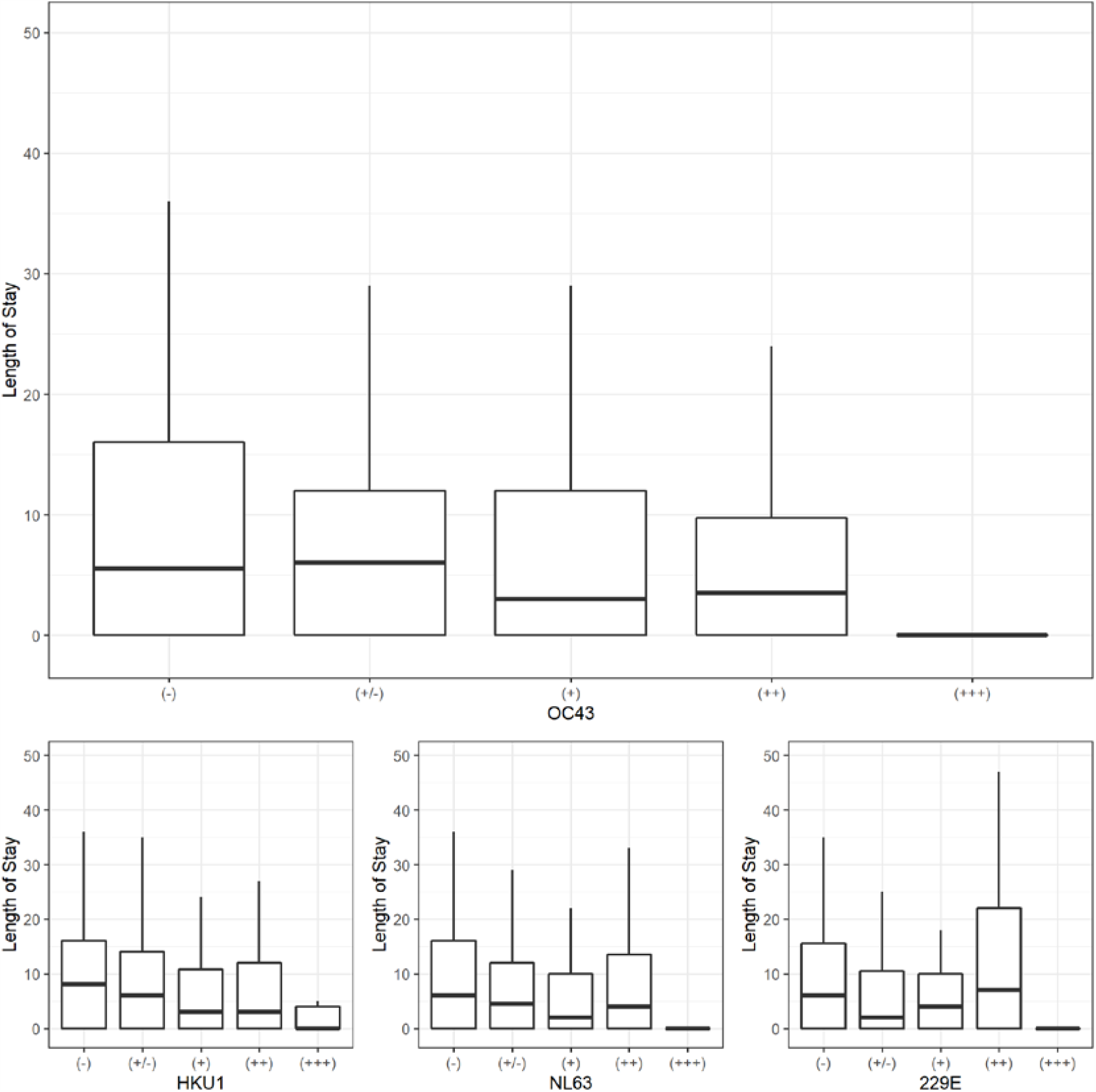
Hospital length of stay in relation to sHCoV antibody level. Patients with very high levels of OC43 antibodies (+++) had significantly shorter LoS than OC43 negative patients (p=0.014). In general, median LoS was shorter for higher antibody levels of OC43 and HKU1.

### Antibody levels against sHCoV in COVID-19 outpatients and inpatients

According to visual determination of band intensities, sHCoV-specific IgG antibodies were less frequently detected in COVID-19 inpatients with critical disease compared to all other COVID-19 patients. Supplemental figure 4 presents a comparison of critical disease (ICU group) with non-critical disease (non-ICU and outpatient groups). This pattern was most pronounced for HCoV OC43 (OC43 antibodies negative: odds ratio for critical disease 1.90 [95% CI 1.08 - 3.32]), followed by HCoV HKU1 (OR 1.32 [95% CI 0.71 - 2.44]), HCoV NL63 (OR 1.10 [95% CI 0.54 - 2.25]) and HCoV 229E (OR 1.20 [95% CI 0.66 - 2.19]). We performed multivariate analysis of the full validation cohort to compare the effect size of OC43 with established risk factors age, sex and BMI. In binary logistic regression, absence of HCoV OC43-specific antibodies (AOR 2.33 [95% CI 1.15 - 4.78]) and male sex (AOR 3.90 [95% CI 2.06 - 7.68]) were the strongest predictors for critical disease. The effect size of OC43 was bigger than age (AOR 1.03 [95% CI 1.01 - 1.05]) and BMI (AOR 1.08 [95% CI 1.02 - 1.16]) in our validation cohort.

Supplemental Figure 5 shows relative sHCoV IgG antibody levels determined densitometrically. COVID-19 patients with critical disease (ICU group) presented with lower sHCoV antibody levels than patients with moderate/severe disease (non-ICU group).

### SARS-CoV-2 antibody levels

Supplemental figure 6 presents results from SARS-CoV-2 IgG antibody measurements in the full validation cohort at first encounter. In general, patients with critical disease had higher SARS-CoV-2 IgG antibody levels compared to moderate/severe inpatients and outpatients. Additionally, IgG seroreactivity against SARS-CoV-2 S1 protein was determined by ELISA (Euroimmun). S1-specific antibody levels detected by ELISA closely matched results of the immunoblot assay (supplemental Figure 7). There was no correlation between NPOC43 antibody levels and SARS CoV-2 S1 reactivity (supplemental Figure 8).

## Discussion

Identification of vulnerable individuals is a key priority in the current stage of the pandemic to guide protective measures and to design vaccination strategies. Recently, we conducted a pilot study comprising 60 patients (10; preprint under review) showing that the presence of IgG antibodies against OC43 NP is associated with a mild course of COVID-19. To corroborate our observations, we conducted a non-interventional validation study with 296 patients from four major tertiary referral centers.

This study clearly confirmed the key finding of the pilot study: Patients with critical COVID-19 requiring ICU treatment showed significantly lower levels of anti-HCoV OC43 NP-specific antibodies compared to non-ICU COVID-19 patients. HCoV OC43 antibody levels were not significantly associated with sex, age or BMI. Accordingly, a lack of HCoV OC43 antibodies can be considered an independent risk factor. The adjusted odds ratio for HCoV OC43 seronegative inpatients to require ICU treatment was 2.68, which might look small at first sight. However, in our validation cohort the risk of HCoV OC43 seronegative inpatients was higher than the risk by increased age or BMI, and lower than the risk by male sex. In accordance with our findings, a recent study based on data from electronic medical records in the Boston area (11) has reported an odds ratio of 0.1 regarding ICU care for SARS-CoV-2 patients with antibodies for endemic coronaviruses.

Cross-reactive humoral and cellular immune responses have been reported to occur among SARS-CoV-2 and other human coronaviruses including OC43 (3, 4, 5, 7, 12, 13). In general, cross-reactivity is expected to be stronger between SARS-CoV-2 and the more closely related seasonal beta-coronaviruses OC43 and HKU1 (14). Since our key finding pertains to OC43 NP IgG-seronegative inpatients, cross-reactivity among seasonal coronaviruses and SARS-CoV-2 does not confound our results. We hypothesize that prior exposure to HCoV OC43 virus facilitates T-cell based and humoral immune responses to SARS CoV-2. While previous work supports this hypothesis (3, 4, 5, 15, 16), further research is needed to gain deeper insights into the underlying immunological processes. According to a recent simulation study (17), cross-immunity between HCoV OC43 and SARS-CoV-2 could affect transmission dynamics of SARS-CoV-2.

Regarding the methodological approach, the immunoblot assay used in our study appears to be only marginally affected by cross-reactivity. Thus, in contrast to SARS-CoV-2-specific IgG levels (15), most patients did not show increases in antibody levels against NP of seasonal coronaviruses, namely OC43 and HKU1 (Figure 1 and supplemental figure 9). Hence, we found no evidence for significant back boosting as described by Shrock et al. (7) regarding our laboratory parameter in this study.

In our validation cohort, COVID-19 outpatients differed in several characteristics as compared to inpatients. In particular, outpatients had more favorable risk profiles with respect to age, sex and BMI. In contrast to the inpatient group, we observed a relatively high variation in sHCoV antibody levels. Prior HCoV OC43 infections may be less important in this group compared to inpatients.

This study has limitations as it was a non-interventional, observational study. We analyzed patient cohorts from major tertiary care referral centers, which could explain the high overall proportion of critical COVID-19 cases. Clearly, a prospective, randomized trial would provide more robust evidence. However, in the current pandemic decisions about vaccination priority and therapy options for COVID-19 inpatients must be taken based on evidence available to date. We observed a consistent pattern in the data from pilot and validation study: HCoV OC43 is the seasonal coronavirus most similar to SARS-CoV-2 (14) and the association measures are stronger for HCoV OC43 than for all other HCoVs.

It is widely accepted that individuals of high age should be vaccinated with priority. If absence of HCoV OC43-specific antibodies conveys more risk than high age, HCoV OC43 seronegative individuals will particularly profit from vaccination and *should be vaccinated with priority*. Approximately 90% of our inpatient cohort were aged 40 years and above, therefore our findings are valid for this age group. For the same reason we propose to determine HCoV OC43-specific IgG antibody levels upon admission of COVID-19 patients to the hospital for individual risk assessment. Table 2 and figure 3 can be used in this context.

## Conclusion

Our results provide evidence that prior infections with sHCoVs, specifically HCoV OC43, can protect against a severe course of COVID-19. Therefore, anti-HCoV OC43 antibodies should be determined in COVID-19 inpatients and considered as part of the risk assessment for individual patients. Hence, we expect individuals tested negative for anti-HCoV OC43 antibodies to particularly benefit from vaccination against SARS-CoV-2, especially with other risk factors prevailing.

## Materials and methods

### Patient cohorts

All patients in the pilot and the validation study experienced SARS-CoV-2 infection as confirmed by RT-qPCR. For each patient, the serum or plasma sample obtained closest to onset of symptoms was examined. Critical disease was defined by invasive ventilation or ECMO therapy in intensive care units; severe disease by oxygen supplementation; moderate disease by hospitalization for other reasons without oxygen treatment; mild disease by limited clinical symptoms (fever, cough, diarrhea, myalgia, anosmia/ageusia) that were managed in an outpatient setting.

In the pilot study, 60 patients were analyzed in the context of the Coronaplasma Project (local ethics committee approval: AZ 2020-220-f-S) and COVID-19 biomarker study (ethics committee approval: AZ 2020-210-f-S) at the University Hospital Münster. This pilot study has enabled us to identify an association between antibody levels against sHCoVs and disease severity of COVID-19.

Figure 5 presents basic demographic information on the validation cohort comprising 296 COVID-19 patients (165 males, 131 females) treated at tertiary care referral centers in Heidelberg, Paris, Erlangen and Regensburg. Inclusion criteria were confirmed COVID-19 diagnosis and age 18 years or above. Five patients suffering from terminal illness were excluded. Overall median age was 60 years (range 18 - 96 years). Median age in the patient group with critical disease (ICU therapy) was 61, with moderate/severe disease (non-ICU) 60.5 and with mild disease (outpatient) 53 years, respectively. There was a significant age difference between these groups (p=0.004); COVID-19 outpatients were on average younger than inpatients.

**Figure 5:**
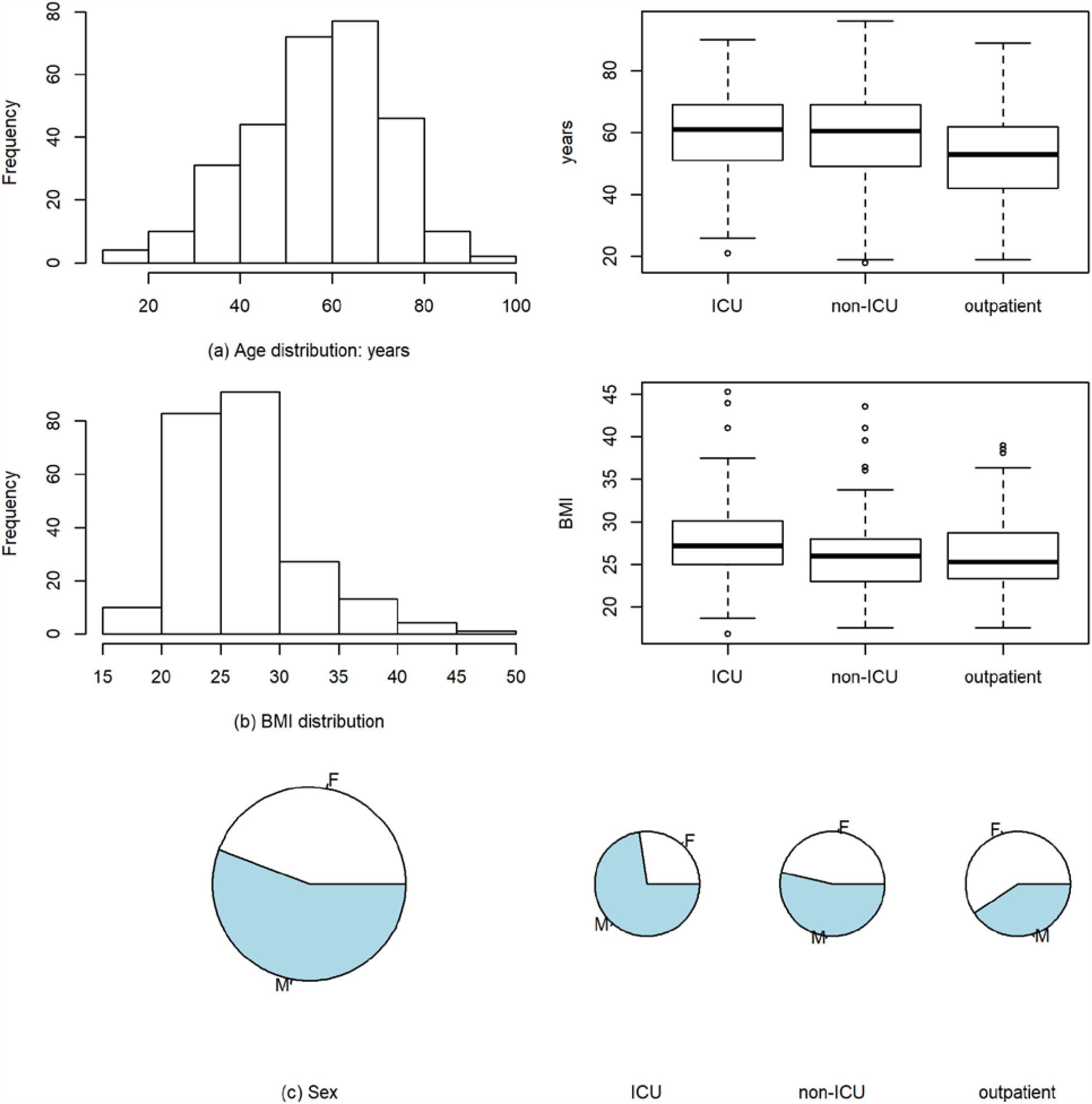
Demographic information on validation cohort (n=296). (a) Age distribution of COVID-19 patients in the validation cohort (global and by type of therapy). (b) BMI distribution. (c) Proportion of males and females: The ICU group comprised more male, the outpatient group more female individuals (p<0.001).

Overall median body mass index (BMI) was 26.0 (range 16.9 - 45.3), in the ICU group 27.2, in the non-ICU group 26.0 and in the outpatient group 25.3. We observed a significant sex difference by type of therapy (p<0.001): While the ICU group comprised more males (77 males, 29 females), the outpatient group included more females (43 males, 63 females). There were 15 fatalities.

The Heidelberg University Hospital contributed 153 COVID-19 patients to the validation cohort (75 males, 78 females). This study was approved by the local ethics committee (ethics committee approval: S-148/2020) and informed consent was obtained from study participants. Enrollment started in March 2020. Median age for patients from Heidelberg was 60 years (range 19-90 years). Forty patients were diagnosed with critical disease, 20 with moderate or severe disease and 93 with mild disease, respectively. Median BMI was 26 (range 17 - 45). Overall, 51 patients showed comorbidities: diabetes mellitus (11), arterial hypertension (12), heart disease (9), hypothyroidism (6), kidney disease (4) and other comorbidities (9).

The second external validation cohort included 49 patients from Assistance Publique-Hôpitaux de Paris enrolled between March and April 2020 in Cochin hospital. All patients were informed and expressed their non-opposition to the study. This non-interventional study was approved by the local ethics committee (AAA-2020-08023). Median age for patients from Paris was 53 years (range 29-80 years). The cohort comprised 22 patients with critical disease, 14 with moderate or severe disease and 13 with mild disease, respectively. Median BMI was 26 (range 18 - 40). Nineteen patients had comorbidities: diabetes mellitus (8), arterial hypertension or cardiovascular disease (6), respiratory diseases (3) and other comorbidities (2).

The third external validation cohort included 46 patients from University Hospital Erlangen enrolled between March and June 2020. This study was approved by the local ethics committee (ethics committee approval: 174_20B) and informed consent was collected from study participants. Median age for patients from Erlangen was 65 years (range 18-96 years). Twelve patients were diagnosed with critical disease, 34 with moderate or severe disease. Overall, 32 patients showed comorbidities, most frequently arterial hypertension (22) and atrial fibrillation (9).

The Regensburg University Hospital contributed 48 patients. The study was approved by the local ethics committee (ethics committee approval: 20-1785-101) and informed consent was obtained from study participants. Median age was 59.5 years (range 19 - 77). 32 patients were diagnosed with critical disease and 16 with moderate or severe disease. 42 patients showed comorbidities, most frequently cardiovascular disease (15) and tumor (8).

### Antibody analyses

Antibody levels against sHCoVs and SARS-CoV-2 were determined with the immunostrip assay recomLine SARS-CoV-2 IgG from Mikrogen GmbH, Neuried, Germany. With respect to sHCoVs, this assay detects IgG antibodies directed against the nucleocapsid protein (NP) of HCoV 229E, NL63, OC43 and HKU1. Of note, this lab test was designed by the manufacturer for high specificity regarding NP-antibodies and does not pick up antibodies against S protein of sHCoVs. For SARS-CoV-2, the assay allows the detection of NP-specific and spike protein (S)-specific antibodies directed against the S1 subunit and the receptor binding domain (RBD). As in the pilot study, analyses were performed at the Institute of Virology/Department of Clinical Virology of the University Hospital Münster according to the manufacturer’s guidelines. To test precision and reliability, internal negative and positive control sera with known antibody reactivity against sHCoVs and SARS-CoV-2, respectively, were included and analyzed as given below. Immunoblot results were analyzed as described in the pilot study (10; preprint under review). In summary, antibody levels were visually determined as ordinal values using the cutoff band of immunstrips as internal reference. Individual coronavirus-specific bands were rated on an ordinal scale as non-detectable (-), below cutoff (+/-), with cutoff intensity (+), above cutoff (++), and very strong intensity (+++), see also supplemental figure 1. Relative antibody levels were quantitatively determined with ImageJ (version 153, 64bit-Version for windows) (18) using the signal intensity of the cutoff band as internal reference (ratio HCoV-specific band to cutoff band). Standardized photographs from immunostrip assays were used for this analysis. Laboratory analyses were performed blinded regarding patient outcome. Quantitative properties of densitometric immunoblot data were validated with a dilution series (supplemental figure 2). In addition, S1-specific IgG antibodies were analyzed by ELISA (Euroimmun, Lübeck, Germany) according to the manufacturer’s instructions.

### Data processing and analysis

Demographical data, type of treatment and length of stay were extracted from the study databases at Heidelberg and Paris. Descriptive statistics and statistical tests were performed with R (version 3.6.1). Ordinal and numerical values were analyzed with exact Wilcoxon test, Kruskal-Wallis test, Chi-squared test, exact Fisher test, Spearman correlation test and binary logistic regression. 95% confidence intervals for odds ratios were calculated with R-package epitools. A two-sided p-value of 0.05 was considered significant.

## Data Availability

For data protection reasons, patient-level data is not available.

## Acknowledgements

We are grateful to Petra Klöters-Plachky, Jutta Mohr, Alina Bauer and Amandine Houvert for excellent technical assistance.

## Author contributions

Dr Dugas, Dr Kühn and Mrs Grote-Westrick had full access to all of the data in the study and take responsibility for the integrity of the data. Dr Tepasse, Dr Vollenberg and Dr Schmidt wrote the study protocol. Dr Dugas, Mrs Grote-Westrick, Dr Tepasse and Dr Kühn contributed equally to this article. Laboratory procedures: Kühn, Grote-Westrick, Lorentzen. Acquisition or interpretation of data: Merle, Tiwari-Heckler, Fontenay, Duchemin, Ellouze, Kremer, Vetter, Fürst, Brix. Concept and design: Dugas, Kühn, Denkinger, Müller-Tidow, Fontenay, Schmidt. Drafting of the manuscript: Dugas, Grote-Westrick, Lorentzen, Kühn, Tepasse. Critical revision of the manuscript for important intellectual content: all co-authors. Statistical analysis: Dugas, Grote-Westrick, Kühn. Obtained funding: Dugas, Müller-Tidow, Kremer.

## Data and material availability

Due to data protection regulations, personal identifiable data cannot be made available.

## References

1. Z. Wu, J.M. McGoogan, Characteristics of and Important Lessons From the Coronavirus Disease 2019 (COVID-19) Outbreak in China: Summary of a Report of 72?314 Cases From the Chinese Center for Disease Control and Prevention. JAMA 2020;323(13):1239–1242. doi:10.1001/jama.2020.2648

2. R.E. Jordan, P. Adab, K.K. Cheng, Covid-19: risk factors for severe disease and death. BMJ. 2020 Mar 26;368:m1198. doi: 10.1136/bmj.m1198

3. A. Grifoni et al., Targets of T Cell Responses to SARS-CoV-2 Coronavirus in Humans with COVID-19 Disease and Unexposed Individuals. Cell. 2020;181(7):1489-1501.e15. doi:10.1016/j.cell.2020.05.015

4. N. Le Bert et al., SARS-CoV-2-specific T cell immunity in cases of COVID-19 and SARS, and uninfected controls. Nature 2020; 584: 457–462. https://doi.org/10.1038/s41586-020-2550-z

5. J. Mateus et al., Selective and cross-reactive SARS-CoV-2 T cell epitopes in unexposed humans. Science. 2020 Oct 2;370(6512):89–94. doi: 10.1126/science.abd3871

6. L. Henss et al., Analysis of humoral immune responses in SARS-CoV-2 infected patients. J Infect Dis. 2020 Oct 31:jiaa680. doi: 10.1093/infdis/jiaa680

7. E. Shrock E et al., Viral epitope profiling of COVID-19 patients reveals cross-reactivity and correlates of severity. Science. 2020 Nov 27;370(6520):eabd4250. doi: 10.1126/science.abd4250. Epub 2020 Sep 29. PMID: 32994364.

8. M. Dugas M et al., Association of contact to small children with mild course of COVID-19. Int J Infect Dis. 2020 Sep 5:S1201-9712(20)30720-7. doi: 10.1016/j.ijid.2020.09.003

9. J.F. Ludvigsson, Systematic review of COVID-19 in children shows milder cases and a better prognosis than adults. Acta Paediatr. 2020;109(6):1088–1095. doi:10.1111/apa.15270

10. M. Dugas et al. Less severe course of COVID-19 is associated with elevated levels of antibodies against seasonal human coronaviruses OC43 and HKU1 (HCoV OC43, HCoV HKU1). preprint: doi: https://doi.org/10.1101/2020.10.12.20211599

11. M. Sagar et al. Recent endemic coronavirus infection is associated with less severe COVID-19. J Clin Invest. 2020 Sep 30:143380. doi: 10.1172/JCI143380

12. Y. Wang et al., Kinetics of viral load and antibody response in relation to COVID-19 severity. J Clin Invest. 2020 Oct 1;130(10):5235–5244. doi: 10.1172/JCI138759

13. K.W. Ng, N. Faulkner, G.H. Cornish et al. Preexisting and de novo humoral immunity to SARS-CoV-2 in humans. Science. 2020;370(6522):1339–1343. doi:10.1126/science.abe1107

14. M. Geldenhuys et al., A metagenomic viral discovery approach identifies potential zoonotic and novel mammalian viruses in Neoromicia bats within South Africa. PLoS One. 2018 Mar 26;13(3):e0194527. doi: 10.1371/journal.pone.0194527

15. P. Doshi, Covid-19: Do many people have pre-existing immunity? BMJ. 2020 Sep 17;370:m3563. doi: 10.1136/bmj.m3563

16. J.J. Guthmiller, P.C. Wilson, Remembering seasonal coronaviruses. Science. 2020 Dec 11;370(6522):1272–1273. doi: 10.1126/science.abf4860

17. S.M. Kissler et al. Projecting the transmission dynamics of SARS-CoV-2 through the postpandemic period. Science. 2020 May 22;368(6493):860–868. doi: 10.1126/science.abb5793

18. ImageJ from NIH. Accessed November 27, 2020. https://imagej.nih.gov/

