## Supplement for "Lack of antibodies against seasonal coronavirus OC43 nucleocapsid protein identifies patients at risk of critical COVID-19"

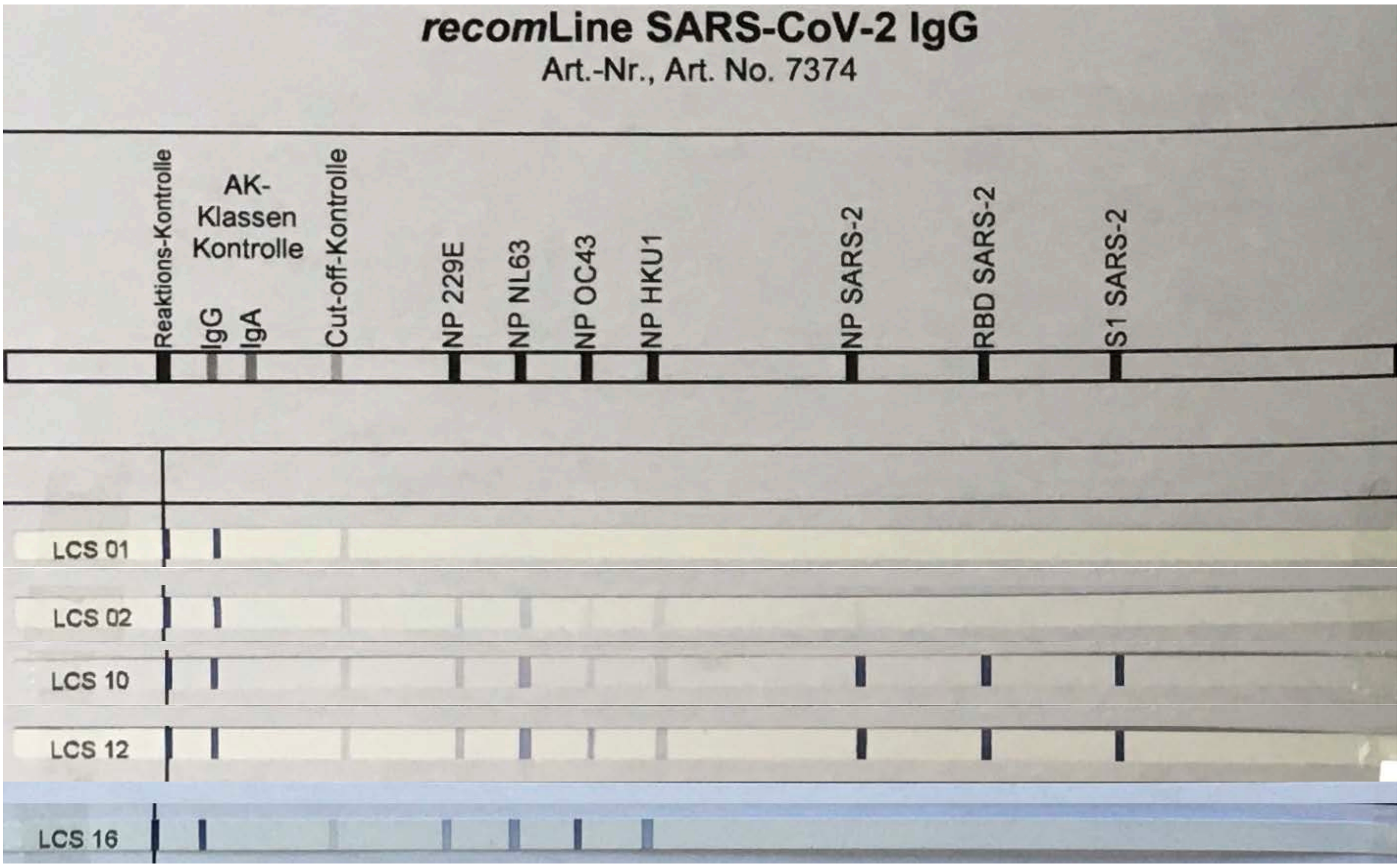


*Supplemental figure 1: Immunoblot with examples of non-detectable (-) [LCS01], below cutoff (+/-) [LCS02], with cutoff intensity (+) [LCS10], above cutoff (++) [LCS12], and very strong intensity (+++) [LCS16] NP OC43 antibodies.*

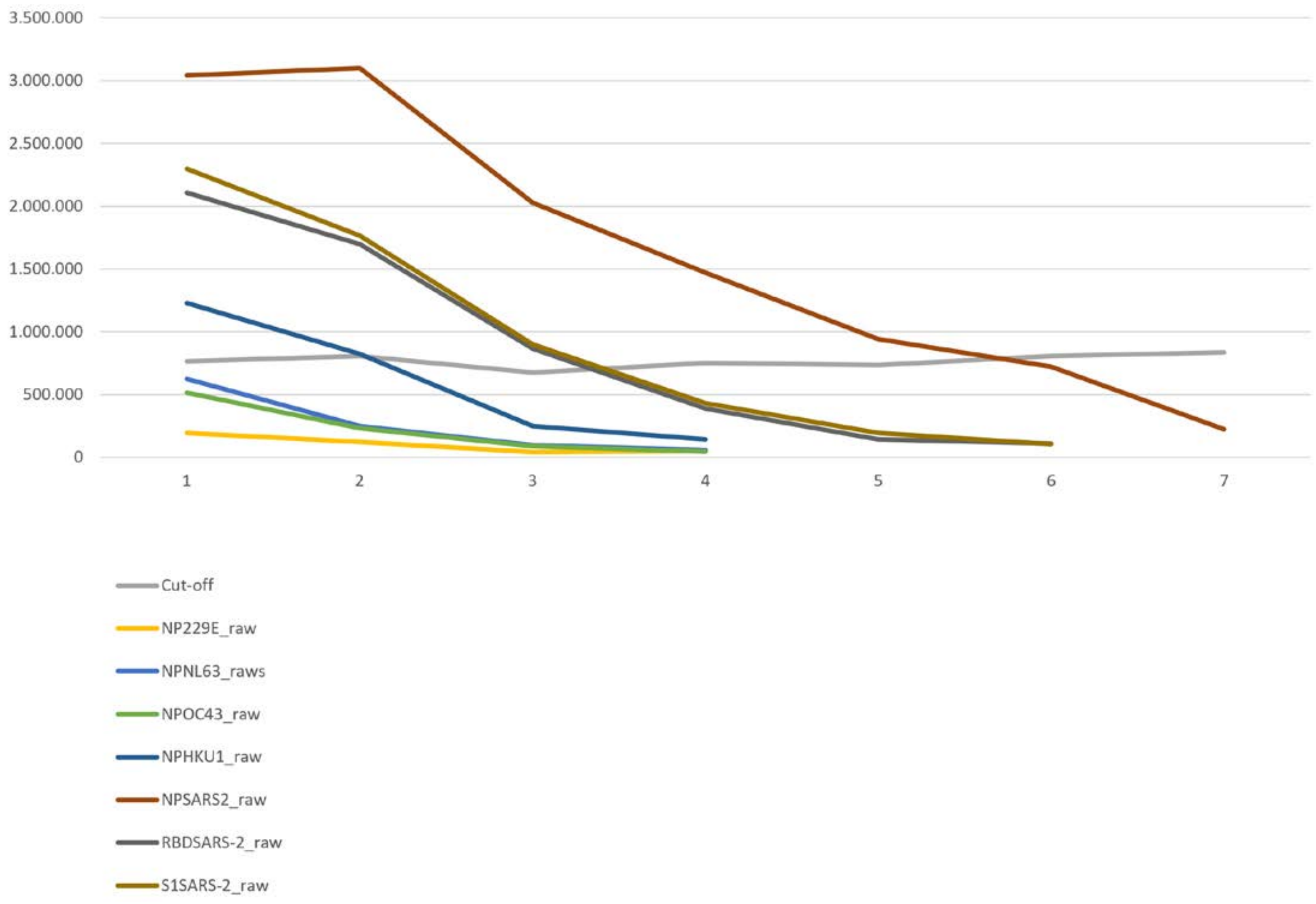


*Supplemental figure 2: Immunoblot dilution series for densitometric antibody measurement. Raw intensity values of 7 different antibodies are presented (NP229E, NPNL63, NPOC43, NPHKU1, NPSARS2, RBDSARS-2, S1SARS-2) for 7 dilution steps. Cut-off level is relatively stable, raw intensity values are approximately linear, therefore densitometric antibody levels can be analyzed quantitatively.*

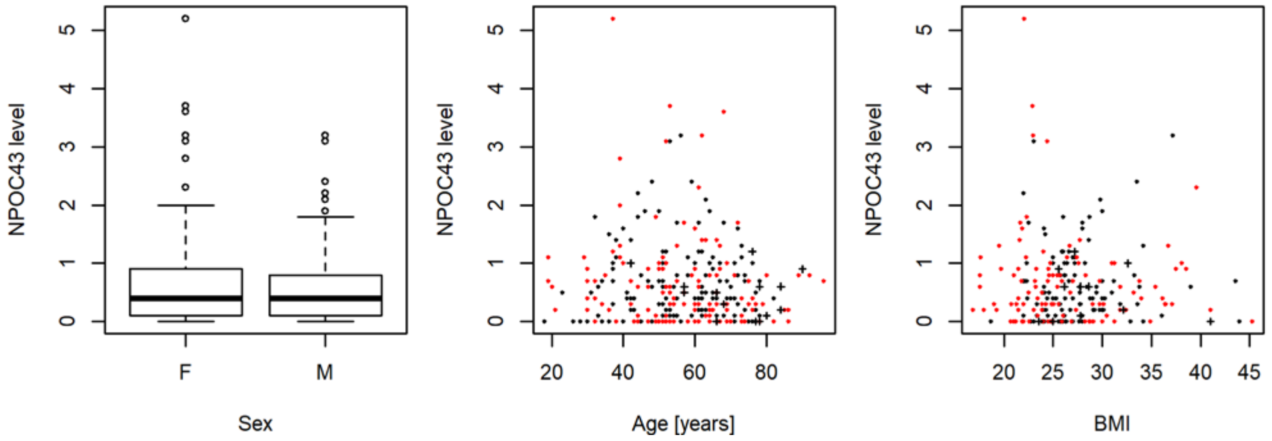


*Supplemental figure 3: Association of HCoV OC43 antibody levels with known risk factors for COVID-19 severity. Black dots indicate male patients, red dots female patients. Crosses denote fatal cases. There was no significant difference regarding HCoV OC43-specifc antibody levels regarding sex, age or BMI in the validation cohort.*

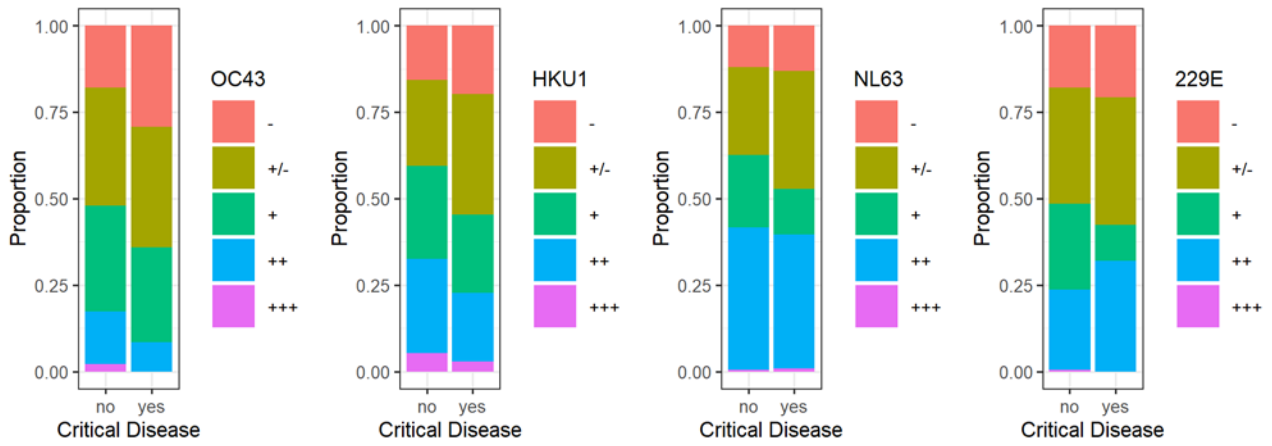


*Supplemental figure 4: Proportion of ordinal HCoV antibody levels from COVID-19 patients with and without critical disease for the full validation cohort (including outpatients). COVID-19 patients with critical disease presented low antibody levels more frequently than patients without critical disease. This difference was most pronounced for HoC OC43 (odds ratio 1.90 [95% CI 1.08 - 3.32]).*

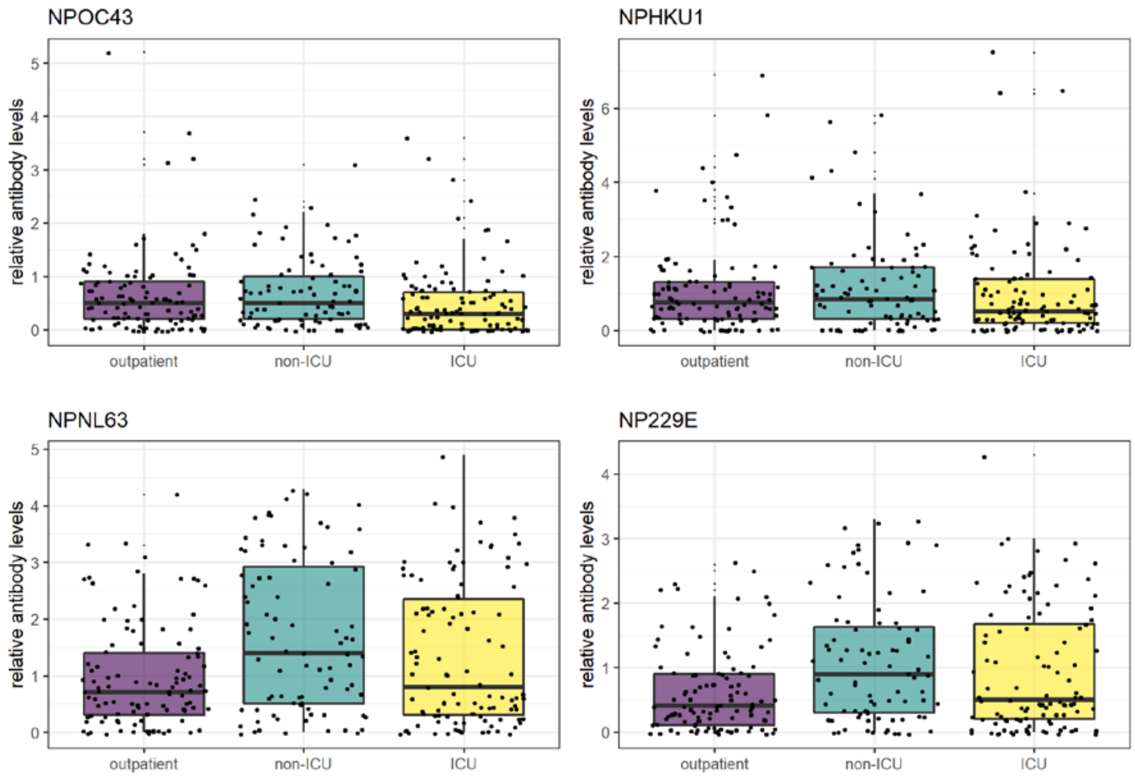


*Supplemental Figure 5: Relative antibody levels against HCoV OC43, HKU1, NL63 and 229E. COVID-19 inpatients from the ICU group presented lower median NPOC43 levels than patients from the non-ICU group (p=0.009).*

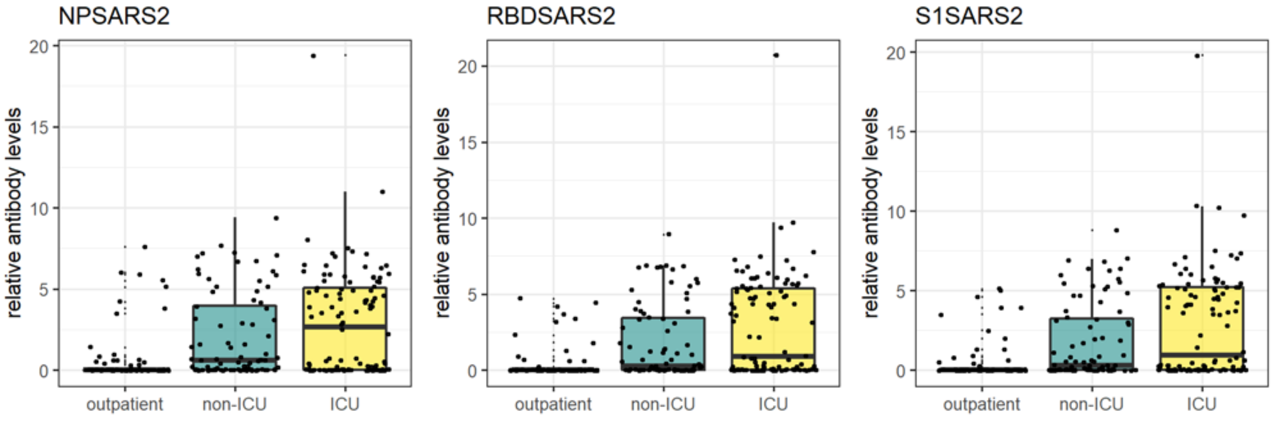


*Supplemental Figure 6: Boxplots of SARS-CoV-2 relative antibody levels for outpatients, patients with severe/moderate disease (non-ICU group) and critical disease (ICU group). Median antibody levels were significantly higher for inpatients (non-ICU/ICU) than for outpatients (p<0.0001) for NP SARS2, RBD SARS2 and S1 SARS2. It has to be taken into account that median time point of sample collection for outpatients was 5 days after symptom onset and 8 days for inpatients, respectively.*

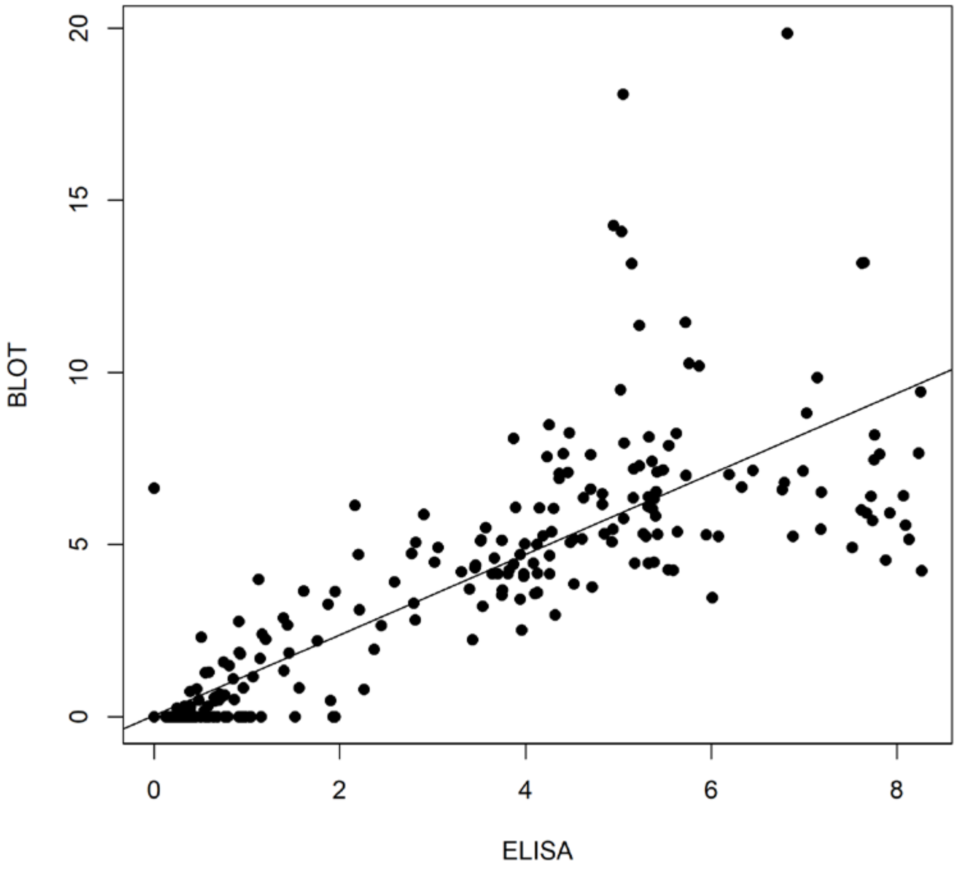


*Supplemental figure 7: Correlation of S1-specific IgG antibody levels by ELISA and Immunoblot. ELISA and Immunoblot values are highly correlated (correlation coefficient 0.84; p<0.001). Therefore antibody levels determined by Immunoblot can be analyzed quantitatively.*

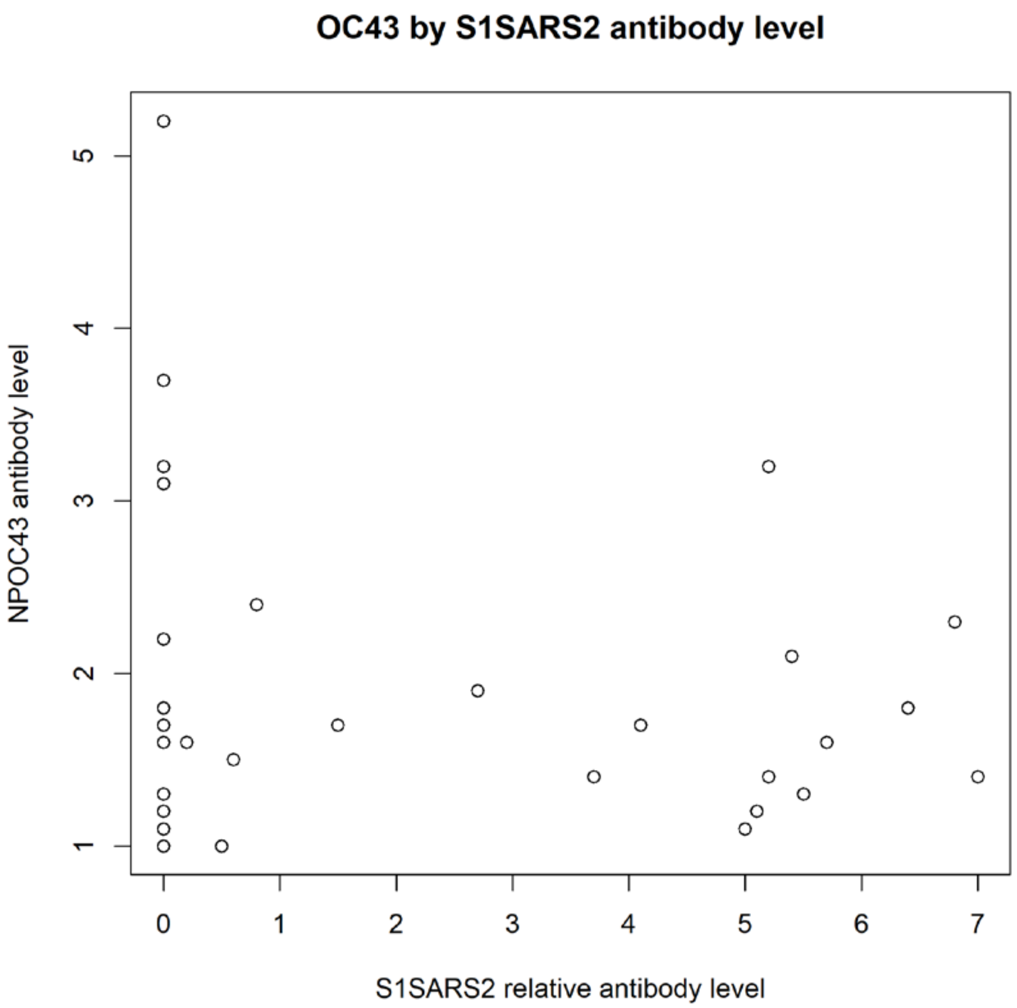


*Supplemental figure 8: Scatterplot of NPOC43 and S1SARS2 antibody levels for patients with NPOC43 antibodies (Immunoblot). There is no significant correlation between NPOC43 and S1SARS2 antibody levels (p=0.33).*

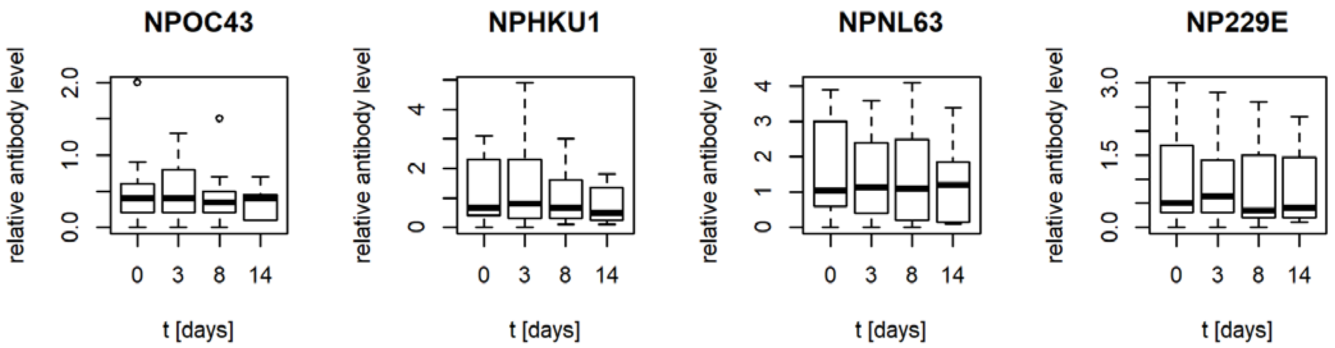


*Supplemental figure 9: In the pilot study, serum samples at several time points (0, 3, 8, 14 days after hospitalization) were available for 14 patients. Regarding the antibody test in this study, there was no evidence for a change of those antibody levels over time (NPOC43: p=0.81; NPHKU1: p=0.93; NPNL63: p=0.98; NP229E: p=0.85).*
